# Risk-Reducing Salpingectomy: Considerations from an OBGYN Perspective

**DOI:** 10.1101/2025.03.11.25323783

**Authors:** Alexandra Lukey, A. Fuchsia Howard, Alice Mei, Michael R. Law, David Huntsman, Celeste Leigh Pearce, Rafael Meza, Gillian E. Hanley

## Abstract

**Background:** With recent evidence that opportunistic salpingectomy is effective in preventing high grade serous carcinoma, it is imperative to consider the optimal use of this procedure. In this research, we investigated the opinions of obstetrician-gynecologists (OBGYNS) about the acceptability of using salpingectomy as a stand-alone surgery for people at higher-than-average lifetime risk (but without a pathogenic variant that increases risk for ovarian cancer) known as ‘risk-reducing salpingectomy’(RRS).

**Methods:** We conducted semi-structured interviews with purposefully sampled practicing OBGYNs in the province of British Columbia, Canada. We used qualitative interpretive description with inductive thematic analysis for data analysis. Our work was informed by the theoretical framework of acceptability.

**Results:** The nineteen participants included physicians from both general obstetrics and gynecology practices, as well as subspecialties. OBGYNs generally found RRS to be acceptable, though this acceptability was conditional on clinical, patient, and system-level factors. Five major themes suggest that: 1) There are risks and benefits of RRS, that if balanced might support acceptability; 2) It is important to define and identify the correct patient for RRS; 3) OBGYNs value patient autonomy in the decision to undergo RRS; 4) Reproductive justice and equity are intertwined and influenced by the history of forced and coerced sterilization; and 5) Formal guidance and the right environment are enablers of RRS.

**Conclusions:** This work provides initial evidence that, from the OBGYN perspective, RRS is acceptable in the right patient population, with considerations from those practicing on the front lines of ovarian cancer prevention.

## Introduction

Ovarian cancer remains a leading cause of cancer deaths in people assigned female at birth, driven by a lack of screening opportunities, late diagnosis, nonspecific symptoms, and the absence of population-based prevention programs.^1,2^ With the current lack of secondary prevention options, attention has turned toward primary prevention interventions such as salpingectomy.^3^ This has been possible due to the science illustrating that HGSC often originates in the fallopian tube.^7,8^ Bilateral salpingectomy, the removal of both fallopian tubes, when undertaken for cancer prevention, is currently practiced opportunistically, either alongside hysterectomy or instead of tubal ligation.^4^ Opportunistic salpingectomy (OS) has been recommended by several international colleges, and its uptake has been increasing.^4–6^ Importantly, OS leaves the ovaries behind, making it a possible prevention opportunity for those at general population level risk for ovarian cancer, as it avoids the increases in all-cause mortality, coronary heart disease, and osteoporosis associated with removal of the ovaries.^6^ Currently, OS is practiced only when a patient is undergoing another pelvic surgery for a different indication, and is thus entirely based on the surgical opportunity and has nothing to do with an individual’s risk for ovarian cancer.

After many decades of study, we have a good understanding of the risk and protective factors for ovarian cancer, which has led to the development of a tool that estimates an individual’s lifetime risk of ovarian cancer. The CanRisk tool has been used to guide risk screening for breast and ovarian cancer since 2007 and has been recommended by several national bodies.^7,8^ The ability to predict an individual’s lifetime risk for ovarian cancer, combined with the considerable safety and effectiveness data that has been generated after 14 years of performing of OS,^9–13^ means that we are in a position to begin considering targeting salpingectomy to individual’s at higher-than-average lifetime risk for ovarian cancer. However, this would not replace the current recommendations for individuals with pathogenic variants that increased their risk for ovarian cancer (i.e. *BRCA1, BRCA2, RAD51C, RAD51D, PALB2* and *BRIP1*). Rather, risk-reducing salpingectomy (RRS), as we are defining it, refers to salpingectomy for the primary indication of cancer prevention, performed as a stand-alone surgery rather than opportunistically for people at moderate risk. While this study will not determine what threshold of lifetime risk will be acceptable, for the remainder of the paper, we will define moderate risk as people who fall below the currently stated thresholds for RRBSO, which is approximately a 5% lifetime risk, but above the average population risk of 1.4%.^14,15^

So far, physician attitudes have been explored regarding opportunistic salpingectomy,^13^ salpingectomy with delayed oophorectomy,^16^ and RRBSO,^17^ but no studies have yet explored physician attitudes toward RRS. This study explored the perceptions of OBGYNs, who would be the main providers of RRS, regarding the acceptability of expanding salpingectomy for ovarian cancer prevention to include RRS. This study explores the acceptability of RRS as well as what considerations would be important for practice from the perspective of OBGYNs.

## Materials and Methods

### Theoretical framing and positionality

As we are interested in the “why” and “how” of OBGYNs’ perceptions of the acceptability of RRS, this work is underpinned by a constructivist research paradigm.^18^ To legitimize and organize this work, we chose to integrate the Theoretical Framework of Acceptability, which includes concepts such as ‘affective attitude,’ ‘ethicality,’ ‘intervention coherence,’ ‘opportunity costs,’ ‘perceived effectiveness,’ and ‘self-efficacy.’^19^ During early data analysis, we added questions informed by the Normalization Process Theory to better explain themes related to “how” RRS would be used in practice. In a constructivist paradigm, the researcher is inseparable from the research process. The primary interviewer (AL) is a PhD candidate and registered nurse with previous experience in qualitative interviewing. The research team is multidisciplinary, with expertise in health policy, ovarian cancer, qualitative methodology, nursing, and health services.

### Study design and setting

We used interpretive description for this study, as this method is suitable for exploratory yet clinically applicable research questions.^20,21^ The products of interpretive description are intended to be applied within a clinical context.^22^ This study took place in British Columbia, Canada which functions as a publicly funded healthcare system that covers medically necessary medical and surgical care. At the time of writing, there are 383 actively practicing OBGYNs serving a population of 5.07 million people. This study was approved by the University of British Columbia Research Board (certificate H22-03281).

### Participant sampling and data collection

We purposefully sampled OBGYNs actively practicing in British Columbia for this study. We excluded any OBGYNs who do not have an active practice license and resident doctors. To ensure practicing status, we cross-referenced the College of Physicians and Surgeons of British Columbia registrant directory. We aimed for maximum variation sampling to gather a variety of viewpoints, including age, number of years in practice, gender, and sub-specialty.^23^ Participants were emailed a letter of invitation to the study. If there was no reply, the study team followed up a maximum of three times to ensure that those who wanted to participate had the opportunity.

We sent out 85 invitations, with 22 OBGYNs consenting to the invitations, although 3 OBGYNs who provided consent did not ultimately complete interviews. When provided, participants cited lack of capacity as a reason for non-participation. We offered a $200 Visa gift card per interview to ensure participants were fairly compensated for their time.

Data collection took place between May and September 2024. Demographic information was collected using a survey hosted on Redcap. All interviews were conducted over videoconference to encourage participation across British Columbia’s diverse geographies and were completed by one author, AL. All interviews were audio-recorded and transcribed verbatim. We used a semi-structured interviewing approach with an interview guide informed by the Theoretical Framework of Acceptability and, later, the Normalization Process Theory (Supplemental Material).^19,24^

### Data analysis

Interpretive description is primarily an inductive methodology.^22^ Therefore, while we used theory in constructing the interview guide, coding was completed using a more inductive data-driven approach, as interpretive description posits that no single theory can encompass the multitude of experiences and realities encountered.^22^ In interpretive description, data analysis and collection happen iteratively. Thus, initial coding began before all the data were collected, and the interview guide was slightly adjusted based on early coding results.

Data analysis was conducted using inductive thematic analysis.^25^ We began coding by immersing ourselves in the data, reading the transcripts, and re-listening to the interviews. Primary coding aimed to capture broad meanings, while secondary coding focused on extracting deeper meanings and connections within the data.^20^ Overall, analysis was performed using a constant comparative process, which involved revisiting codes and data. Themes were discussed within the study team (AL, FH, GH). Data management and coding were completed using NVivo^TM^ version 12.^26^

## Results

Our sample of 19 OBGYNs included participants from general obstetrics and gynecology practices, as well as sub-specialties such as gynecologic oncology, reconstructive surgery, endometriosis, and menopause (Table 1). Participants had practice experience ranging from 4 to 40 years, and the majority were women (13/19). OBGYNs generally perceived RRS to be acceptable, though this acceptability was conditional on clinical, patient, and system-level factors. Five main themes described important considerations that informed OBGYN’s perspectives on the acceptability of RRS: 1) There are risks and benefits of RRS that, if balanced, suggest potential acceptability; 2) It is important to define and identify the correct patient for RRS; 3) OBGYNs value patient autonomy in the decision to undergo RRS; 4) Reproductive justice and equity are intertwined and influenced by the history of forced and coerced sterilization; and 5) Formal guidance and the right environment are enablers of RRS. These themes are described below.

### 1. There are risks and benefits of RRS, that if balanced suggest potential acceptability

OBGYN commentary suggested that the acceptability of RRS was primarily driven by their strong desire to prevent ovarian cancer and the physician’s belief in the efficacy of salpingectomy. Despite the high degree of comfort performing the surgery and belief in the efficacy of salpingectomy, nearly all OBGYNs brought up concerns related to complications of RRS. Interviewees discussed the importance of factoring the risk of surgical complications, including patients’ surgical history and comorbidities, into decisions about surgery, particularly in the case of elective surgery. Several participants also expressed concern that patients don’t always fully grasp the real risks of elective surgeries, which would be a concern if RRS was offered. “*I think people don’t really recognize that there are risks of this type of surgery*.” (Participant 1156)

OBGYNs also brought up considerations related to healthcare resource management that impacted how they perceived the acceptability of RRS as a purely preventative surgery. This contrasted with opportunistic salpingectomy, which one OBGYN described: “*I usually like to do a two-hit wonder, right? So, if we’re taking out tubes, I want to see that there are other benefits.*” (Participant 1154) Participants lauded the benefits of opportunistic salpingectomy, whether it is for another surgery to improve quality of life, contraception, gender-affirming care, or expanding into other specialties such as colorectal surgery.

Concerns about limited access to operating room time to perform RRS were also highlighted in relation to healthcare resources, as stated by one OBGYN, “*It’s just so bad up here that I can’t reconcile how I could ever even in the next 10 years justify offering that without the evidence and without a guideline saying that I should*.” In contrast, several surgeons discussed the potential savings from a healthcare system perspective. “*It will improve the system level when you consider the context… The resources that are put into treating women with advanced ovarian cancer and the outcomes that you achieve*.” (Participant 1171) When discussing the risk of complications of RRS, one OBGYN said: “*I think it’s a risk that would be warranted to prevent a very deadly cancer. So, for me, it’s mostly about the system capacity. That’s my main concern*.” (Participant 1172)

Most OBGYNs agreed that RRS made sense when the lifetime risk was high enough and without other contraindications to surgery. Specifically, surgeons expressed interest in knowing the number needed to treat to ensure RRS is worthwhile from a healthcare system perspective. *“The numbers are important because there are many other cancers that are more prevalent. But we’re not necessarily removing those [organs]”* (Participant 1160). Overall, OBGYNs discussed the importance of RRS outweighing risks both at a patient and a healthcare system level.

### 2. It is important to define and identify the correct patient for RRS

The importance of identifying people who would benefit from RRS and the task of finding the “moderate-risk” individuals were challenges identified by the OBGYNs. Opportunities to offer RRS were broadly divided into two categories: first, people who are proactively seeking ovarian cancer prevention, such as those who may have already requested genetic screening, had another cancer, or were socially aware of ovarian cancer; or, second, people who would somehow be “screened” into being offered RRS.

Multiple OBGYNs discussed people who were already in contact with the cancer system in some way. Regarding previous cancer patients, one OBGYN said, *“When they finish their treatment, that’s when they start to think, ‘Okay, what else can I do to stay safe and healthy?’…That’s when they start to ask about tubes and ovaries coming out.”* (Participant 1151) Another group that OBGYNs brought up as candidates was people who had received negative genetic testing results but still wanted to take steps to prevent cancer, or those with risk factors who wanted genetic testing but did not meet the criteria for publicly funded testing. *“If they did not meet hereditary cancer program referral criteria for testing. I would still feel comfortable offering them the surgery.”* (Participant 1172)

History-based screening was the other method the OBGYNs indicated as a way of offering RRS to the right patients. The overwhelming majority of OBGYNs brought up family history of ovarian cancer as a flag to consider RRS. *“I would think strong family histories probably do warrant it. It just would come down to how and when these people interact with the care system.”* (Participant 1151) Several OBGYNs highlighted that a strong family history, in the absence of known genetic variants, was qualitatively enough to raise the lifetime risk of ovarian cancer to a threshold to offer RRS if the patient was otherwise a good surgical candidate who wanted the surgery. *“I think really, if there’s a patient with any family history of ovarian cancer or any family history of early breast cancer, then that patient should be having a discussion with an OBGYN.”* (Participant 1166)

### 3. OBGYNs value patient autonomy in the decision to undergo RRS

Some OBGYNs described their prior experience offering RRS when patients requested it, often due to an awareness of the risks associated with ovarian cancer or relevant aspects of their “social history,” described by OBGYNs as having someone in their social circle, family member or otherwise who had experienced a diagnosis of ovarian cancer. OBGYNs commonly emphasized the importance of informed consent, allowing patients to decide for themselves.

> “We have quite a lot of knowledge about doing it [salpingectomy] and the safety and risks of it as an elective procedure. So, that information I have, so, I can educate them. Okay, this is the risk of this procedure in an elective setting. And because we’re already doing that for other indications, it’s up to them to decide what indication is really acceptable for them.” (Participant 1162)

OBGYNs highlighted the variability in patients’ risk tolerance, noting how different patients respond and relate to perceived risk based on their individual experiences. Generally, participants described a desire to support patient decision-making unless the OBGYN considered the surgical risk truly too great. Participants also generally expressed high confidence in their ability to have discussions and allow patients to weigh benefits and risks. “*I think gynecologists are used to having these conversations all the time. Because, unless you’re in gyne-onc, we are not doing these surgeries to save people’s lives. We are doing it to improve quality of life*.” (Participant 1164)

OBGYNs described their reluctance to engage in medical paternalism but also expressed uncertainty about how they would approach cases where younger patients might request RRS and the concern over regret for RRS. A couple of the OBGYNs indicated that in such instances, they would be inclined to broach alternative options and recommend people go on the birth control pill if ovarian cancer was the main concern because it’s reversible. “*I guess there are other ways of approaching it, like you can go on the birth control pill that can reduce your risk of ovarian cancer. And guess what, if you don’t want to stay on the pill, you can stop it. If I take your tubes out, that’s going to reduce your risk. But if you want them back in, that’s going to be a problem*” (Participant 1160). There were varied opinions—some thought young people could be offered RRS, but others felt there should be guidelines for providers.

### 4. Reproductive justice and equity are intertwined and influenced by the history of forced and coerced sterilization

Several OBGYNs raised concerns about Canada’s history of forced and coerced sterilization, particularly in the context of RRS being not only a cancer prevention intervention but also a form of permanent sterilization. The participants who discussed this issue also described feeling uncertain about how to ensure cultural safety when offering RRS, especially regarding Indigenous patients. One OBGYN emphasized the difference in situations where a patient requests RRS versus one where it is initially offered by the physician, “*It’s different than them coming to us saying ‘my family’s complete, I’d like to have sterilization’ versus me saying to them ‘hey, can I offer you this sterilization procedure?*’” (Participant 1158) Several participants also discussed communication barriers as an equity concern with offering patients RRS.

Another participant raised concern about potential misuse in a fee-for-service healthcare environment where physicians would be paid for performing RRS: “*You know, who’s promoting it? This could go sideways very, very quickly if healthcare providers were promoting an intervention for which they get paid*.” They expanded on their point: “*Who’s driving the bus? I think this is one area where the last people that should be driving the bus are those people that would be doing the procedures*” (Participant 1171).

On the other hand, OBGYNs also brought up issues of inequity in not offering RRS. One participant, described their concerns related to inequities in how RRS might be taken up. This participant emphasized that those with high health literacy and privilege tend to receive the best preventative care, and there is a significant risk of this disparity becoming further entrenched with RRS unless addressed thoughtfully.

> “[We need to] make sure that we’re including women of low socioeconomic status and lowest education levels. Because again, the patients that have usually come requesting things given their family history, it’s usually Caucasian, highly educated women. And of course, if patients bring it up, and it’s a concern, I’m happy to address it. But I’m not offering this to my patients that don’t speak English and don’t know their family history. So I think for those women to get included from the beginning, it’s important. Yeah, if it’s going to become a population-level intervention potentially down the road.” (Participant 1145)

### 5. Formal guidance and the right environment are enablers of RRS

While nearly all participants described opportunistic salpingectomy as part of their regular practice, and several described already practicing RRS as defined in this paper, participants highlighted that there is no formal guidance for risk reduction for people at increased risk of ovarian cancer but who do not meet current criteria for RRBSO and who do not have another indication for surgery. Guidance, such as in the form of national guidelines, was suggested by participants regarding who would be a candidate for RRS, similar to what currently exists for RRBSO for individuals with high-risk polygenic variants. One participant highlighted the need for guidelines and the reassurance this would provide physicians when offering surgery, “*you’ll give us the green light to go on certain people, and so that’ll give us the confidence to do it on certain people and all the people who are at that medium risk*.” (Participant 1161)

Participants also discussed the importance of collaboration with primary care. Several participants brought up the example of the cervical cancer prevention program, with clear referral criteria for primary care on when to refer to specialist care. Ovarian cancer does not have “one” clear referral criteria, although many discussed family histories as a driving factor.

However, OBGYNs indicated that more guidance would be needed for consistent care. As described by one participant, “*Often patients tell us, ‘Oh, you know, I have this high history of ovarian cancer in my family.’ And you’re like, ‘Okay, is one sister enough? Is one grandmother enough?*’” (Participant 1145)

Gaps in primary care were also often discussed as a potential barrier to the equitable delivery of RRS. Participants highlighted the issue of the lack of patient attachment to primary care providers.

> “I think the bigger problem is that 30% of people have no long-term healthcare association. So like family practice would be identifying them. There has to be, you know, a reasonable risk discussion that this is an option because you’re now a moderate-risk individual, and so I think it starts from the bottom, which is basically the primary care providers have to be educated.” (Participant 1154)

Conversely, OBGYNs often praised the culture of practice change in BC and discussed pride in BC as the birthplace of salpingectomy. Many also warned that the culture in BC related to salpingectomy is not widespread, commenting on more hesitancy within Canada. One OBGYN reflected on their willingness to change practice: “*Do we want to be the 1st adopter? Or do we want to be early adopters? But definitely not a late laggard, you know. It’s very interesting to think about. Where do you want to be in that pendulum of change?”* (Participant 1172)

## Discussion

Our study found that RRS was acceptable to OBGYNs in BC, but only when certain patient and resource considerations were met. Specifically, OBGYNs were interested in the number needed to treat to ensure that RRS was coherent from a public payer system perspective, and in making sure that eligibility criteria were clear to ensure that the benefits of RRS outweighed the risks of surgery for individual patients. They were also focused on supporting autonomy for their patients. Additionally, they discussed the need for formal guidance, such as national guidelines, especially as public awareness of salpingectomy grows, and more patients are requesting salpingectomy alone for ovarian cancer prevention. The need for tools and guidance to effectively partner with primary care providers was also emphasized.

Current guidelines suggest that salpingo-oophorectomy should be considered for individuals with a lifetime risk of ovarian cancer of 5% or greater, and after the age of 35.^14,15^ OBGYNs in our study agreed that a strong family history is a sufficient reason to consider RRS. They also expressed comfort in offering salpingectomy alone as a risk-reduction procedure below the 5% threshold. In line with NICE guidelines, which call for increased research into accurately assessing cancer risk, OBGYNs were uncertain about specific cut-offs related to family history, particularly given the uncertainty surrounding less common pathogenic variants and their relation to family history.^14^ Similarly, NICE guidelines suggest that people with as low as a 2% likelihood of carrying a pathogenic variant should be offered genetic counseling.^14^

While RRS was deemed acceptable in terms of OBGYN affective attitudes, coherence, and perceived effectiveness, embedding RRS into practice may be inhibited by factors outside the scope of OBGYN practice, according to Normalization Process Theory.^24^ For instance, participants in our study discussed the lack of attachment to primary care providers as a barrier to patients receiving preventive care. In areas where not all patients have equal access to primary care, multiple levels of inequity must be addressed to ensure access to risk-reducing salpingectomy.^27^ Additionally, OBGYNs in some settings raised concerns about limited operating room availability, similar to the issues reported by Gelderblom et al.^13^ Therefore, acceptability was influenced by the opportunity cost construct of the Theoretical Framework of Acceptability, particularly in settings such as Canada where healthcare resources are constrained.

From the OBGYN perspective, once thresholds are established from a system perspective, they thought there would be little issue with enrollment. As described by Normalization Process Theory, enrollment occurs when individuals work together to participate in a new initiative or practice.^24^ Several OBGYNs pointed to the uptake of opportunistic salpingectomy as a case study demonstrating the ability of OBGYNs to change practice when presented with sufficient evidence.

As highlighted by our participants, future work on RRS should prioritize equity and cultural safety from the outset.^28^ It has already been shown that opportunistic salpingectomy is offered inequitably across different demographic groups and care settings.^29,30^ Therefore, guidelines and shared decision-making tools should be developed through a cultural safety and trauma-informed lens.^31^

### Strengths and Limitations

Our study has several limitations. First, while the goal of qualitative research is not to be universally generalizable, our work may be at risk of response bias, as it likely attracted participants who already find salpingectomy acceptable and “early adopters” who were more willing to participate. We sought to mitigate this through purposive sampling. However, the context of this study may have influenced the sample’s awareness of and support for salpingectomy as a method of ovarian cancer prevention. Therefore, the results may not fully represent the opinions of all BC OBGYNs, nor those in the wider Canadian or international gynecology communities.

## Conclusions

This research provides initial evidence that RRS is acceptable from the OBGYN perspective, including considerations from those practicing on the front lines of ovarian cancer prevention. Our findings indicate that more research is needed to determine and understand optimal and acceptable thresholds for the recommended lifetime risk, both to better counsel patients and to justify this intervention from a health system perspective. Future research should focus on ensuring there is equity in access to RRS, as well as to better understand the patient perspective regarding RRS.

## Supporting information

Interview guide

## Declarations

### Ethics Approval/Consent for participation and publication

This study was approved by the University of British Columbia Research Ethics Board (certificate H22-03281). All participants completed consent to participate in this research and consent for publication including use of de-identified direct quotes.

### Competing Interests

The authors declare that they have no competing interests

### Funding

Canadian Institutes of Health Research, Canada Graduate Scholarships — Doctoral program. North Family Foundation, Vancouver General Hospital & University of British Columbia Hospital Foundation-Gynecologic Cancer Initiative Clinical Trials Group Accelerating Grants Program. The funders of this research had no role in the study design; in the collection, analysis, and interpretation of data; in the writing of the report; or in the decision to submit the article for publication.

### Author Contributions

AL, MRL, DH, CLP, and GEH were responsible for funding acquisition. AL, FAH, MRL and GEH were responsible for the design of the study. AL, FAH, and GEH conducted the data analysis. AL drafted the manuscript, and FAH, AM, MRL, DH, CLP, RM, and GEH read and critically revised the paper. All authors approved the final manuscript.

## Data Availability

The data generated in this study are not publicly available due the risk of re-identification, but are available upon reasonable request from the corresponding author for study finding verification purposes.

## Acknowledgements

We would like to acknowledge all the physician participants who took the time to complete interviews and make this research possible.

## Table Legend

Table 1. Sample characteristics of participants

## Notes

### Competing Interest Statement

The authors have declared no competing interest.

### Author Declarations

Ethics committee/IRB of the University of British Columbia gave ethical approval for this work

