## Supplementary material for "Risk-Reducing Salpingectomy: Considerations from an OBGYN Perspective": Interview guide

**Acceptability of Risk-Reducing Salpingectomy: ARISE Study**

**Interview Guide (OBGYNs)**

- What does your current practice look like regarding counselling patients about salpingectomy?
- You indicated you think would do risk reducing salpingectomy at [insert lifetime risk] could you explain your thinking about that?
- How do you think an individual’s lifetime risk of ovarian cancer ought to be factored into recommending salpingectomy? For instance, should we be using a tool like the CanRisk calculator? Or what information would you need to know to counsel the patients about risk reducing salpingectomy?
- What factors would you weigh most heavily if you were to recommend risk-reducing salpingectomy?
- What are your main concerns when thinking about offering risk-reducing salpingectomy to patients?
- If risk reducing salpingectomy were to become part of standard practice, how do you think these moderate risk people should be identified?
- Another part of this research is looking at whether we can use population-based data to identify these moderate risk people. For instance the data collected by the ministry of health to create a risk prediction algorithm to flag patients who may be at higher risk. How would you feel about patients being identified this way?
- Previous clinicians we’ve talked to have indicated that having someone with expertise in having conversations about salpingectomy is important. What do you think might be important to consider for these conversations?
- Is there anything that I haven’t asked you about that you think is important to talk about?
