## Supplementary material for "Risk-Reducing Salpingectomy: Considerations from an OBGYN Perspective": Table 1

*Table 1. Sample characteristics of participants*

| *Characteristics* | *Overall (n =19)* |
| --- | --- |
| Practice years (mean (SD)) | 16.8 (10.3) |
| Gender (%) |  |
| Woman | 13 (68.4) |
| Man | 6 (31.6) |
| Specialty training (%) |  |
| Endometriosis, Pelvic Pain and Advanced Laparoscopic Surgery | 2 (10.5) |
| Vulvovaginal Disorders/ Sexual Medicine | 1 (5.3) |
| Gynecologic Oncology | 3 (15.8) |
| Pediatric and Adolescent Gynecology | 1 (5.3) |
| Maternal Fetal Medicine | 2 (10.5) |
| Menopause | 1 (5.3) |
| Practice Setting |  |
| Private office or clinic | 14 (73.7) |
| Academic health sciences centre | 8 (42.1) |
| Community Hospital | 10 (52.6) |
| Non-academic teaching hospital | 3 (15.8) |
| Telemedicine | 3 (15.8) |
